# *in utero* HIV exposure and the early nutritional environment influence neurodevelopment in infants before age three: findings from an evidenced review and meta-analysis

**DOI:** 10.1101/2020.08.27.20182774

**Authors:** Marina White, Kristin L. Connor

## Abstract

The developing brain is especially vulnerable to infection and suboptimal nutrition during the pre- and early postnatal periods. Exposure to maternal HIV infection and antiretroviral therapies (ART) *in utero* and during breastfeeding can adversely influence infant (neuro)developmental trajectories. How early life nutrition may be optimised to improve neurodevelopmental outcomes for infants who are HIV/ART-exposed has not been well characterised. We conducted an up-to-date evidence review and meta-analysis on the influence of HIV exposure *in utero* and during breastfeeding, and early life nutrition, on infant neurodevelopmental outcomes before age three. We report that exposure to maternal HIV infection/ART may adversely influence expressive language development, in particular, and these effects may be detectable within the first three years of life. Further, while male infants may be especially vulnerable to HIV/ART exposure, few studies overall reported sex-comparisons, and whether there are sex-dependent effects of HIV exposure on neurodevelopment remains a critical knowledge gap to fill. Lastly, early life nutrition interventions, including daily maternal multivitamin supplementation during the perinatal period, may improve neurodevelopmental outcomes for infants who are HIV-exposed. Our findings suggest that the early nutritional environment may be leveraged to improve early neurodevelopmental trajectories in infants who have been exposed to HIV *in utero*. A clear understanding of how this environment should be optimised is key for developing targeted nutrition interventions during critical developmental periods in order to mitigate adverse outcome later in life, and should be a priority of future research.

## Introduction

In pregnancy, HIV infection has profound effects on maternal physiology, fetoplacental development, and pregnancy outcomes. Globally, targeted public health programmes and the increasing availability of antiretroviral therapies (ART) provided 85% of 1.3 million pregnant women living with HIV access to the treatments necessary to prevent mother-to-child transmission (PMTCT) in 2019^1^. As a result, the number of children born annually who are HIV-exposed *(in utero* and during breastfeeding) but are uninfected (HEU) themselves is rising, and it is estimated that there are currently 15.2 million children globally who are HEU^2^. The extent to which exposure to maternal HIV infection and ART may have lasting impacts on the development of children who are HEU, and the influence of other decisive exposures, including early life nutrition, on these outcomes remains to be thoroughly explored. This gap in understanding limits the development and use of early interventions to improve neurodevelopmental trajectories, tailored to support developmental susceptibilities that children who are exposed to HIV *in utero* or during breastfeeding may experience.

Importantly, persistent immune dysfunction and inflammation experienced by pregnant women on ART^3^ may heighten the risk of neurodevelopmental disorders in offspring^4^. Circulating levels of maternal inflammatory markers during the prenatal period directly associate with the organization of neural networks in the newborn brain, most significantly in regions critical for attentive abilities, social behaviour, communication and motor coordination, and are predictive of working memory abilities at two years of age^5^. The developing brain is also vulnerable to the effects of suboptimal maternal nutrition, as the fetal and neonatal brain depend on nutrition supplied by the mother, prenatally through transplacental transfer, and postnatally through breastfeeding and other enteral feeds, to support rapid brain development during these critical periods^6^. Breastfeeding is recommended for women living with HIV who are on ART, particularly where undernutrition, diarrhea and pneumonia are common causes of infant mortality^7^, and breastfeeding is associated with reduced hospitalization in infants who are HEU during their first year of life^8^. It is critical to understand whether breastfeeding may be beneficial for neurodevelopment in infants who are HEU, as this could provide a target for early nutrition-related interventions.

It is also necessary to consider interactions between exposure to infectious disease (such as HIV) and malnutrition, given that these exposures often coexist in socially inequitable contexts. For example, individuals living with HIV are vulnerable to food insecurity as a result of decreased economic capacity, and food insecurity has been associated with reduced care access and poorer clinical outcomes for people living with HIV^9^. Maternal immunosuppression related to HIV infection may also be exacerbated by malnutrition^10^, and the comorbidity of these exposures for infants *in utero* and during the breastfeeding period may be more detrimental than the occurrence of one of these circumstances alone. The multiple intersections of HIV/AIDS and food insecurity have led to calls for integrated nutrition and HIV/AIDS programming^11^.

Previous reviews have reported poorer neurodevelopmental outcomes in school-aged children who are HIV-exposed and perinatally infected (HEI)^12,13^ and HEU may have persistant, negative effects on neurodevelopment until at least age eight^14^. However, to our knowledge, there has been no review and meta-analysis of evidence related to early life nutritional exposures and neurodevelopmental outcomes in infants who are HEI or HEU. Nutritional factors are likely important determinants contributing to these outcomes, as vitamin A and maconutrient supplementation have been linked to reduced mortality and improved growth outcomes, respectively, among children who are HEI^15^, and nutritional status is a critical contributor to early neurodevelopment^6^. Successful prenatal interventions to improve infant development by addressing nutrition access have included folic acid, calcium and multivitamin supplementation, while vitamin A supplementation and promotion of exclusive breastfeeding have proved effective postnatally^16^. Similar early nutritive interventions may prove effective for rising number of infants who are born annually HEU, and who may be even more vulnerable to the programming effects suboptimal nutrition *in utero* and postnatally^2^. An improved understanding of these relationships is key to optimising early interventions for maximal, positive impact on neurodevelopment and function^6^, allowing children to thrive.

Here, we aimed to answer how, and to what extent, do HIV exposure and early life nutritional factors during critical windows of brain development influence infant neurodevelopmental outcomes. Specifically, we first synthesised evidence on how exposure to maternal HIV infection and ART *in utero* and during breastfeeding affects the neurodevelopmental outcomes of infants who are HEI or HEU in the first three years of life. Next, we investigated how early life nutritional exposures (breastfeeding practices, nutrition-related interventions and food security circumstances) may modify the developmental trajectories of these infants. We also examined sex differences in neurodevelopment and how early life nutrition factors may influence these outcomes for infants who were exposed to HIV, given that male infants are often more susceptible to developmental insults experienced *in utero* in comparison to female infants^17^.

## Methods

### Inclusion criteria

Article screening took place as part of a larger scoping review, inclusive of papers relating to growth, immune, and neurodevelopmental outcomes in infants who are HEI or HEU, and the influence of early life nutritional factors on these outcomes. PRISMA reporting guidelines were followed (Supplementary Table 1)^18^. Within the neurodevelopment theme, eligible study designs were randomised controlled trials (RCTs), controlled clinical trials, cohort, case series, case-control, or cross-sectional studies. Articles that included at least one group of infants exposed to maternal HIV infection (either HEI, HEU or HIV-exposed but infant infection status unknown) who had a neurodevelopmental assessment before three years of age were eligible for inclusion. This age criteria allowed us to capture information on early life development within a relatively focused window, inclusive of the recommended period of exclusive (six months) and mixed-breastfeeding (24 months)^19,20^. The first 36 months (3 years) of life are also an especially sensitive period for neurodevelopment, as the brain’s structure and functional capacity rapidly develop during this time^6,21^. Detecting differences in developmental outcomes before three years of age is critical for determining whether children who are HEI or HEU may benefit from the introduction of additional support during critical developmental periods, and in what areas the support is needed, in order to improve developmental trajectories. Lastly, articles that reported on data collected prior to 2000, when international PMTCT efforts were first launched^22^, were excluded from review in an effort to increase comparability across studies and relevance to the current-day context of treatment and management of HIV infection in pregnancy, given the drastic shifts in the global response to HIV over the last 20 years.

### Information sources and search terms

PubMed, CINAHL, ProQuest, and Web of Science were used to retrieve peer-reviewed publications on pre-defined key terms (Supplementary Figure 1) to extract papers related to growth, neurodevelopment, and immunological status in infants exposed to maternal HIV infection *in utero* or during breastfeeding. The search yielded a total of 20642 peer-reviewed articles in the English Language, including 16501 duplicates which were subsequently excluded (EndNote Web), leaving 4141 articles eligible for level one screening.

### Article screening and data collection

#### Level one: screening for growth, neurodevelopment and immune outcomes

A three-level screening process was constructed with the inclusion and exclusion parameters set to capture relevant articles (Supplementary Figure 2). References for 4141 articles from the EndNote Library were exported into the Distiller SR software for systematic review. At level one, article titles were reviewed for relevance and classified according to theme (growth, neurodevelopment, or immunological status), resulting in the exclusion 2858 articles. These largely included review articles, articles on policies to prevent mother-to-child transmission of HIV, the socio-cultural impacts of living with HIV, or counselling for mothers living with HIV on breastfeeding practices. Articles that discussed policies on infant vaccination schedules and administration, or studies that strictly reported on a country’s mortality, morbidity, and survival trends were also excluded. Where a clear assessment of eligibility based on the article title was not possible, the article was carried forward to level two screening.

#### Level two: screening for neurodevelopment

In the second level of screening, references for 358 articles related to neurodevelopment and 183 that remained unclassified were extracted for abstract review. At this level, 447 articles were excluded where neurodevelopmental outcomes were not reported in the study, or assessments had not occurred prior to three years of age.

#### Level three: screening and data collection for the neurodevelopmental theme

The third level screening included full article review and data collection, for which 94 articles within the neurodevelopmental theme were included. Pre-structured forms within the literature review software captured neurodevelopmental outcomes related to cognition, motor, behaviour, language, and neurostructural development. At this level, articles that indicated a primary exposure of interest other than maternal HIV infection or early life nutritional factors, and that did not report comparisons of neurodevelopmental outcomes based on either of these factors, were excluded. These studies included RCTs investigating relationships between timing of ART initiation or different ART and infant health outcomes, and observational studies reporting on the influence of child-caregiver interactions on infant development among infants exposed to HIV, as these were considered outside of the scope of this review. Lastly, given the time lapse between the initial literature search (during November 2016) and the write up of this review, an additional search was performed by hand on March 25^th^, 2020 using the same set of pre-determined key words in each of the four databases to capture any relevant studies published since the original screen. 11 additional articles were identified in this secondary search and in total, 24 articles met full eligibility criteria and were included for evidence review and meta-analysis.

#### Screening for early life nutritional factors within the neurodevelopmental theme

All 24 articles that met the primary inclusion criteria for assessment of neurodevelopmental outcomes were subsequently screened for inclusion of data related to maternal nutritional status, breastfeeding practices or reports of food insecurity during pregnancy and the postpartum period, or infant nutritional status in the first three years of life, for which nine articles met at least one of these criterion.

#### Methodological quality assessment

Articles were assessed for methodological quality according to study design using the following scales: Newcastle-Ottawa Quality Assessment scale^23^, the Quality Appraisal Tool for Case Series (18-item checklist)^24^, and the Cochrane Collaboration’s Tool for Assessing Risk of Bias^25^ for cohort studies (n=15), case series (n=6), and RCTs (n=3), respectively, as has been previously recommended^26^. Criteria for methodological quality assessment were set a priori for each scale and are described in detail in Supplementary Table 2. In brief, for cohort studies, comparability of exposed and non-exposed cohorts was determined based on whether or not analyses controlled for infant sex and age. Notable differences in neurodevelopment and vulnerability to insult have been recorded for male versus female infants^27^, and variation in age at neurodevelopmental assessment between groups was considered a potential confounder^28^. Where neurodevelopmental data were longitudinal, adequacy of follow up cohorts was determined when subjects lost to follow up were minimal (set at <20%), or analyses were run to establish similarity between infants retained at follow up vs. not, as previously recommended^28^. For case series, characteristics of the cohort that were important to report were pre-defined as: number of participants (infants), age range of infants with neurodevelopmental assessments at each time point, and infant sex. Intervention and co-intervention definitions were modified to be exposure of interest (maternal HIV infection and ART) and co-exposure of interest (infant HIV and treatment status). For RCTs, “other bias” was defined as an assessment of participant compliance to intervention. While methodological quality was not a primary outcome of interest in this review, it was deemed necessary to help inform our interpretations and weighing of results across studies.

### Data analysis

A random effects meta-analysis was performed on data from studies that used the Bayley Scales of Infant Development-3^rd^ ed. (BSID-III)^29^ and reported scaled or composite scores for the cognitive sub-scale^30^. A random effects model was chosen because it considers between-study variance^31^. However, it is recommended to have a minimum of five studies when using a random effects model^31^, and only three studies reported BSID-III scaled or composite scores for the gross and fine motor, and expressive and receptive language sub-scales. Thus, as we were underpowered to synthesize the results for these four subscales through meta-analyses, we report their combined effect estimates for information’s sake only and discuss these findings qualitatively. Raw scores were not considered in meta-analyses, as they are not age-adjusted. Between-group comparisons for infants who were HEU and HIV-unexposed, uninfected (HUU) were considered. Hedge’s *g* was chosen as an estimate for effect size measurements as it has been shown to be accurate in the case of small sample sizes^32^. Heterogeneity (I^2^) was not calculated as it has shown to be highly biased in small sample sizes^33^. Statistical significance was confirmed at α=0.05 and results are presented as Hedge’s *g* (95% confidence interval).

## Results

### Study location, demographics and design

The articles under review included participant data from 17 countries (Supplementary Figure 3). Of the cohorts included in studies under review, 57% (n=17) were from Africa, followed by 17% (n=5) from North America, 13% (n=4) from South America, and 7% (n=2) from both Asia and Europe. One article reported on data from cohorts in Brazil, Argentina, Peru, Mexico, Bahamas, and Jamaica^34^. South African cohorts had the highest representation in studies under review (29%, n=7).

For the 24 studies included for review, cohort characteristics, including study groups, age at neurodevelopmental assessment, timeline of neurodevelopmental assessments (cross-sectional or longitudinal) and outcome themes are reported in Figures 1 and 2 in adapted Graphical Overview for Evidence Reviews (GOfER) diagrams^35^. Studies comparing neurodevelopmental outcomes based on infant HIV exposure status (primary inclusion criteria, n=15) are summarised in Figure 1, and studies that reported on both early life nutrition-related variables and infant neurodevelopment (primary and secondary criteria, n=9) are summarised in Figure 2. Within the 15 studies that met the primary inclusion criteria only, there were six that reported on longitudinal neurodevelopmental outcomes (for cognitive [n=3], motor [n=4], language [n=1] and neurostructural [n=1] themes) for infants before age three^34,36-40^, and nine that reported cross-sectional data (for cognitive [n=8], motor [n=8], language [n=4], behavioural [n=4] and neurostructural themes [n=2])^41-49^. For the nine studies that included analyses on early life nutrition-related variables and infant neurodevelopment, cohort characteristics and comparison groups based on nutritional intervention (if relevant) are reported in Figure 2. Within these nine studies, two reported data on longitudinal neurodevelopmental assessments (for cognitive [n=2], motor [n=2] and language [n=1] outcomes)^50,51^, and seven reported cross-sectional assessment data (for cognitive [n=7], motor [n=7], language [n=7] and behavioural [n=3] outcomes)^52-58^.

**Figure 1.**
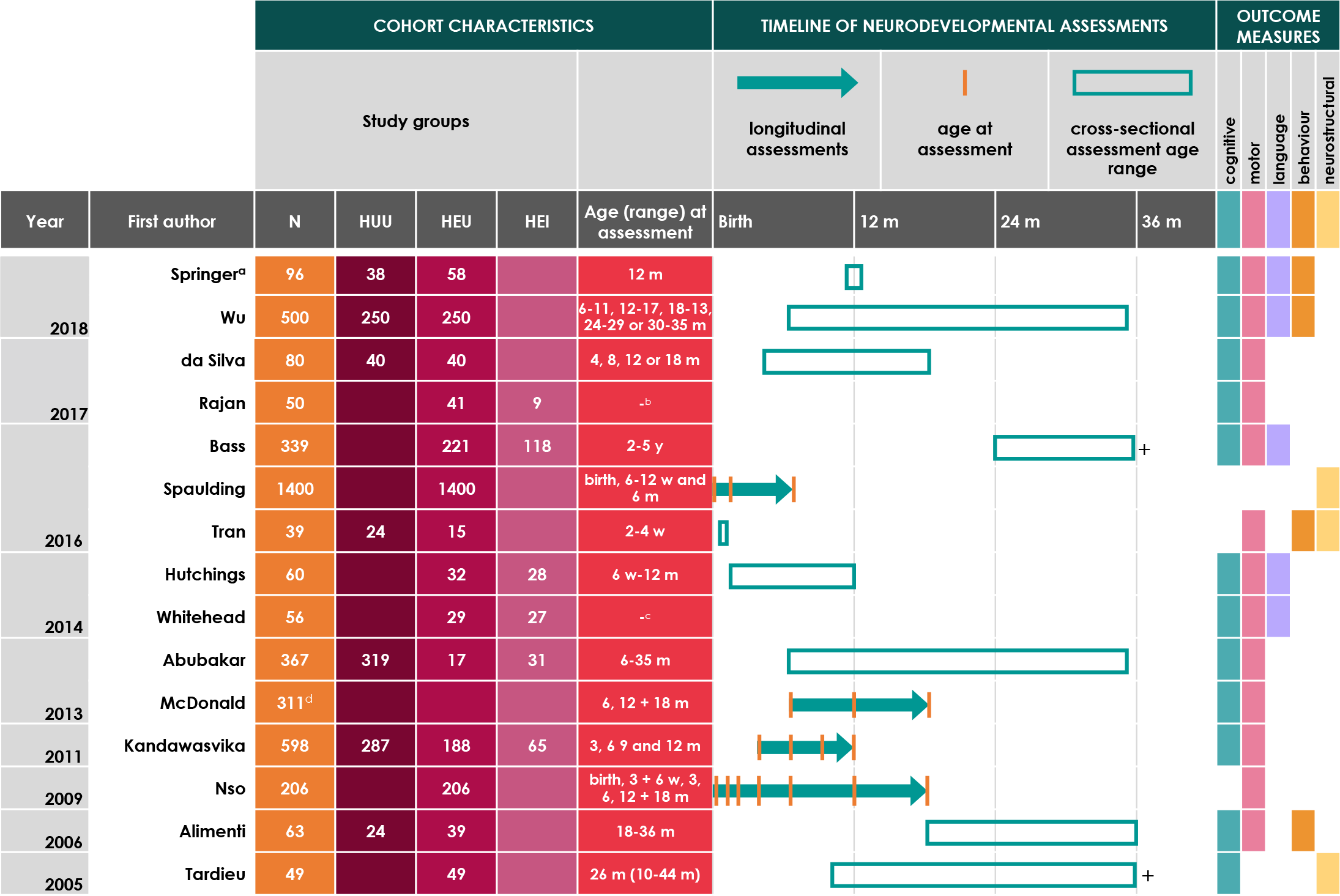
Adapted Graphical Overview for Evidence Reviews (GOfER) Diagram^35^ of studies reporting on neurodevelopmental outcomes in infants in relation to HIV-exposure status. ‘+’ indicates that assessments were performed for infants beyond 36 months of age. HUU = HIV-unexposed, uninfected; HEU = HIV-exposed, uninfected; HEI = HIV-exposed, infected; d = days; w = weeks; m = months; y = years. ^a^Two studies report on data from the Mother and Infant Health Study (MIHS) cohort^49,53^. ^b^Development was assessed for all children at enrolment (range 6-18 m) and after 3 months. A third assessment was done for 25 children after 6 months of enrolment. ^c^Infants under one year of age were eligible for recruitment. Neurodevelopment was assessed at baseline (prior to initiating ART for infants who were HEI) and again three and six months later. Breakdown of infant ages at baseline, second and third assessments was not provided. ^d^A breakdown according to infant HIV status for the 311 infants who had neurodevelopmental assessments not available. All were HIV-exposed. 139 HEI and 519 HEU assessments were used in analysis for both cognitive and motor outcomes (repeat measures for infants were included). Data from this infant cohort is reported in another study under review^51^.

**Figure 2.**
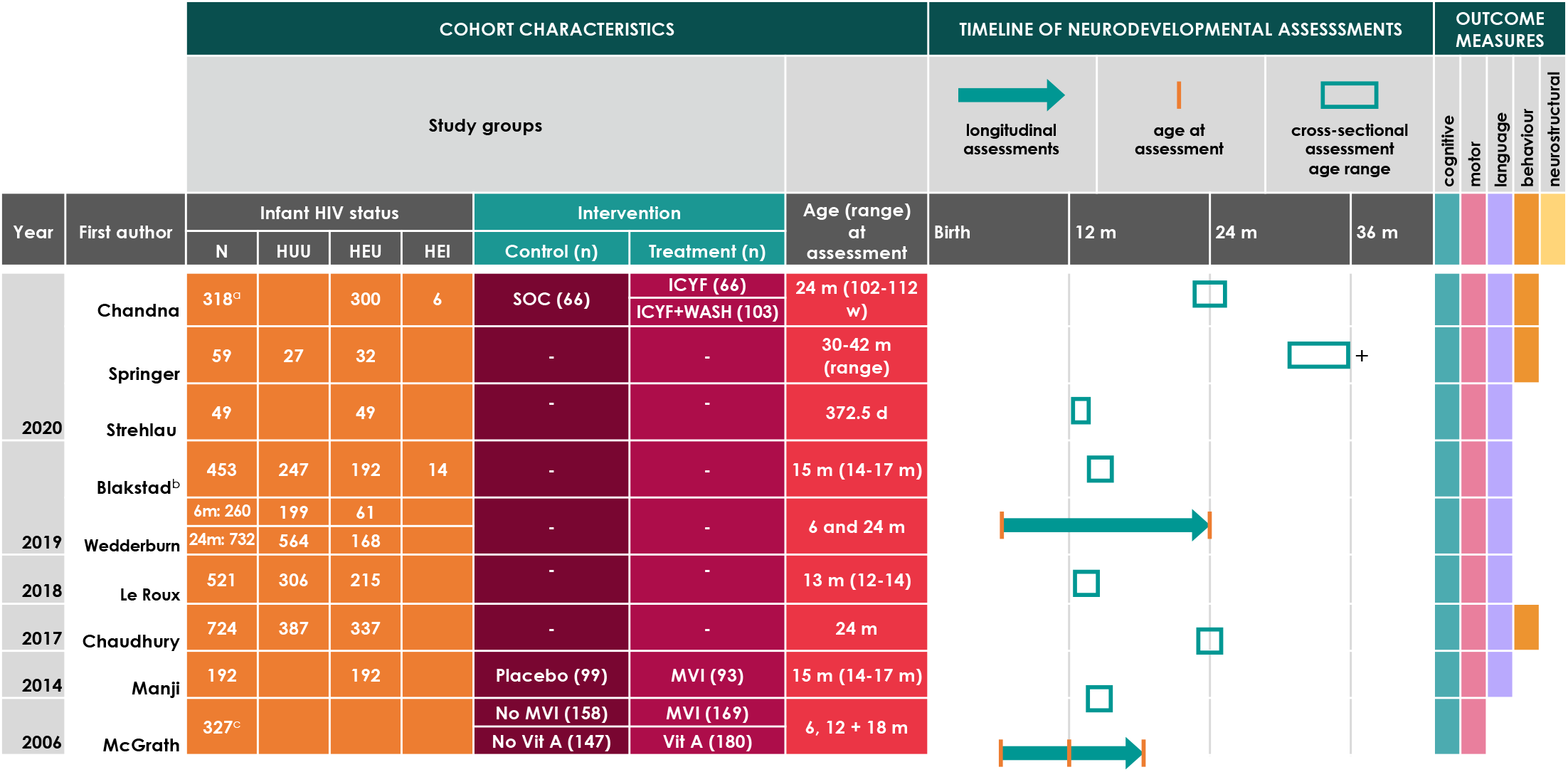
Adapted Graphical Overview for Evidence Reviews (GOfER) Diagram^35^ of studies reporting on early life nutritional factors and neurodevelopmental outcomes in infants affected by exposure to maternal HIV infection. Detailed descriptions of the study treatments or interventions are presented elsewhere (Supplementary table 3). HUU = HIV-unexposed, uninfected; HEU = HIV-exposed, uninfected; HEI = HIV-exposed, infected; SOC = Standard-of-care; ICYF = Infant and young child feeding; WASH = Water, sanitation and hygiene; d = days; w = weeks; m = months. ^a^There were 12 infants included in the analyses whose HIV status was unknown. ^b^Reports on two infant cohorts who were enrolled in two separate micronutrient trials in Tanzania. One of these cohorts is reported on in another study under review^58^. ^b^AII infants were HIV-exposed. Authors did not report cohort breakdown according to infant HIV status but did report that infant HIV status did not modify relationships between multivitamin supplementation and neurodevelopmental outcomes.

### Study measurement tools

The measurement tools employed to assess neurodevelopmental outcomes are reported in Tables 1 and 2 for each study. The most common assessment scales were the Bayley Scales of Infant Development—1^st^ to 3^rd^ editions^29,59,60^. To assess neurostructural outcomes, two articles used structural imaging techniques (magnetic resonance imaging and diffusion tensor imaging^44,48^ and one article used the World Health Organization standardised growth measures^01^ to ascertain microcephaly in infants^34^).

**Table 1.**
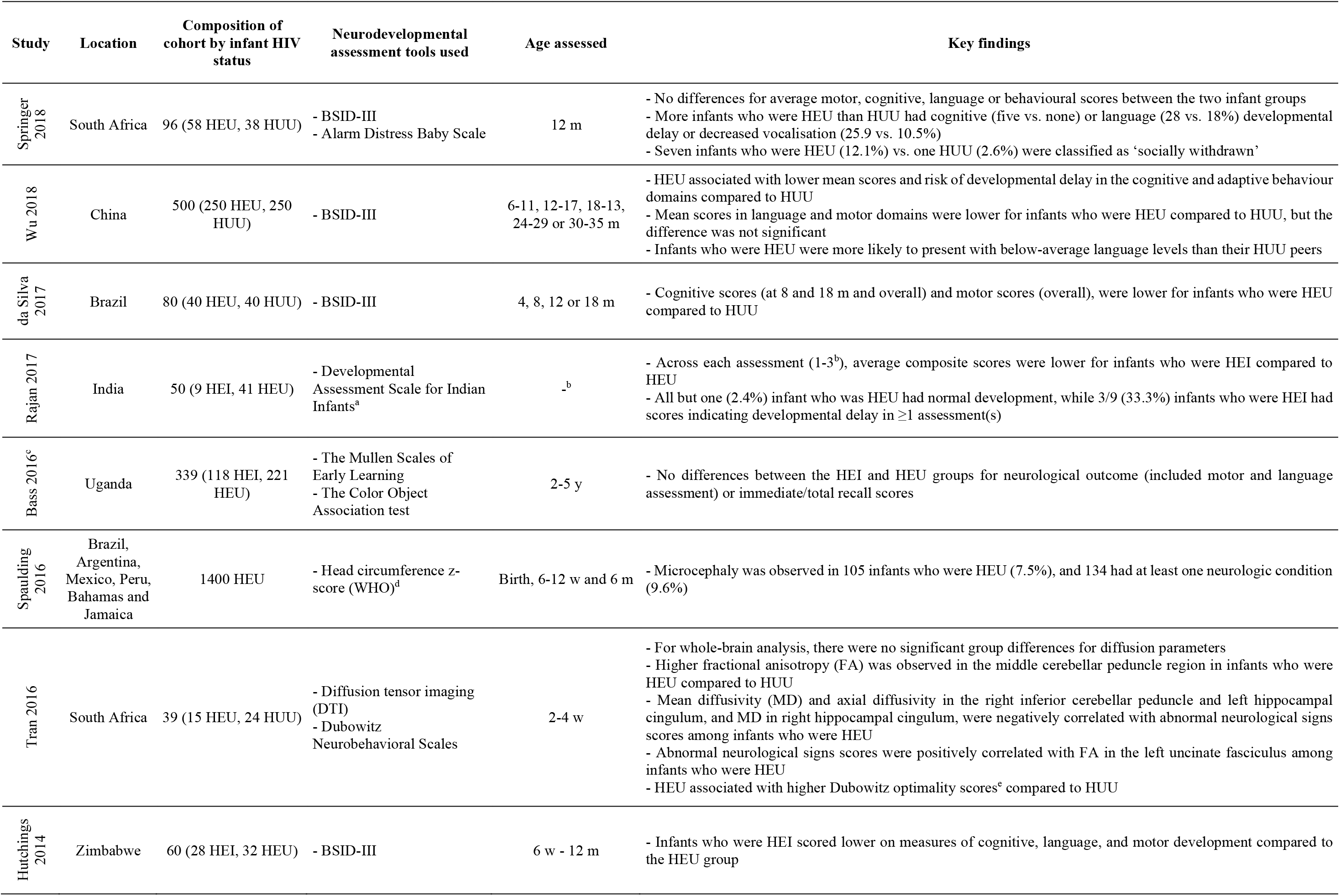
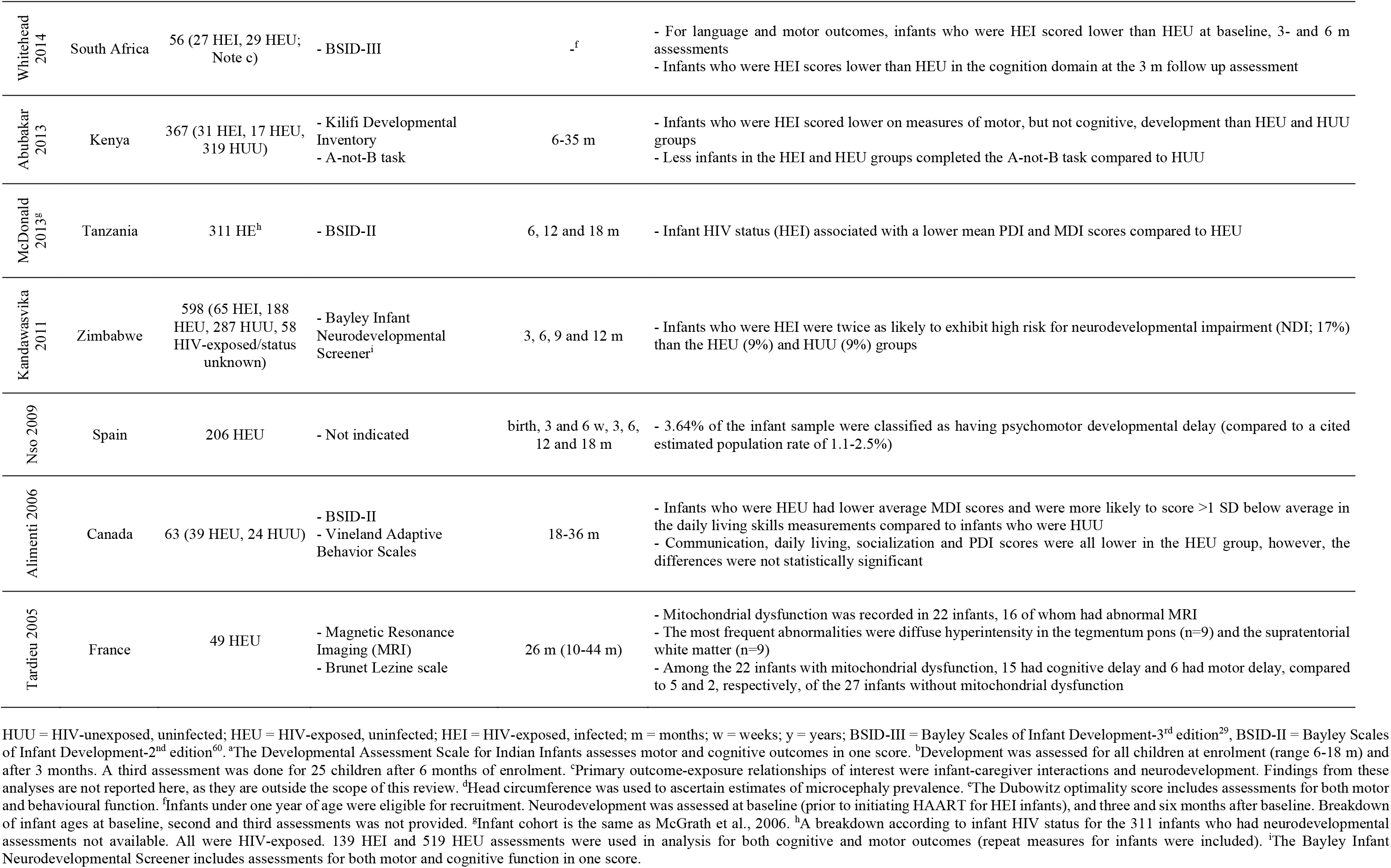
Summary of key findings on neurodevelopmental outcomes from birth to 36 months of age in infants born to mothers living with HIV.

**Table 2.**
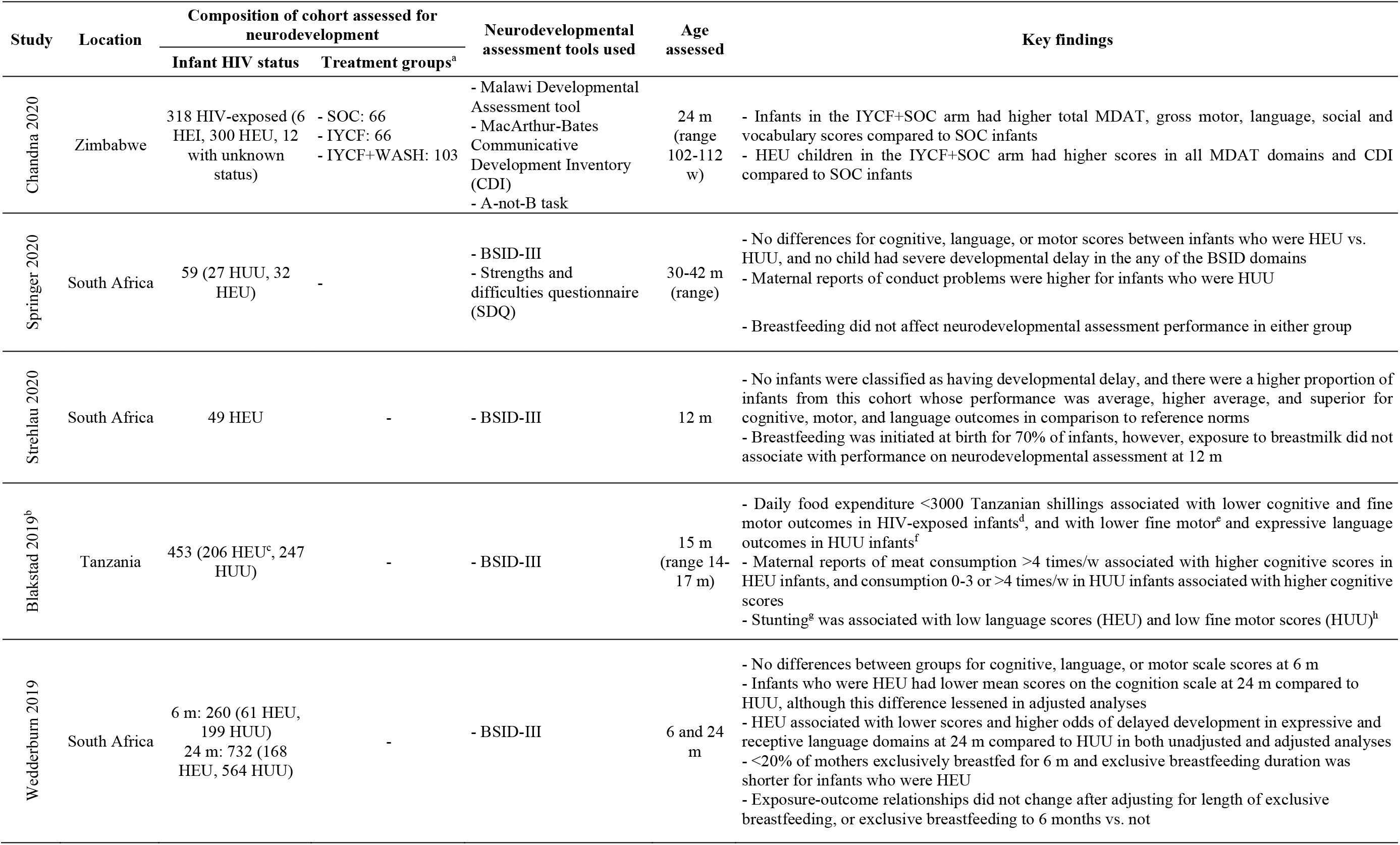
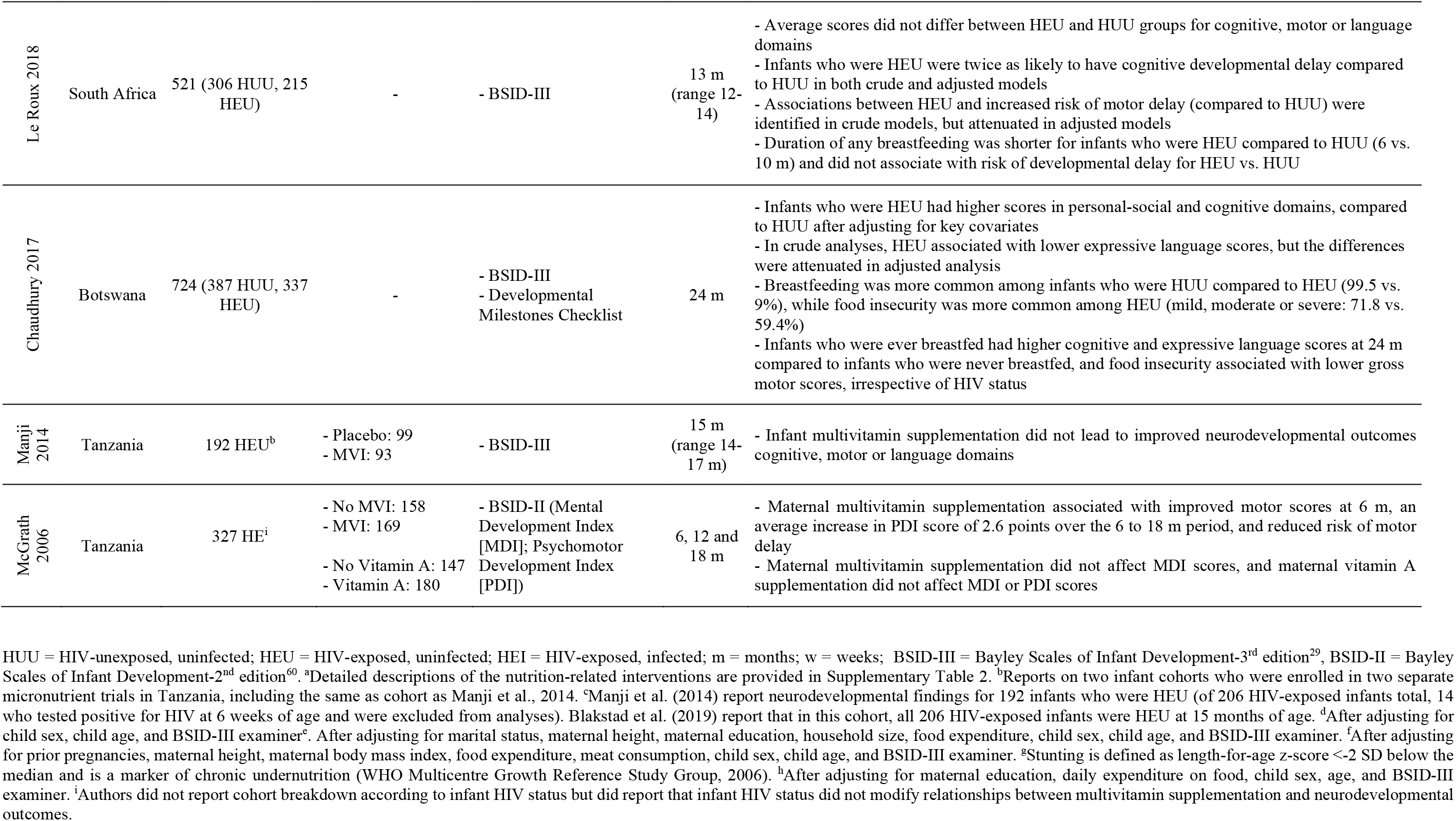
Summary of key findings on the influence of nutrition-related factors and interventions on neurodevelopmental outcomes in infants born to mothers living with HIV.

### Methodological quality assessments

Among the cohort studies under review, assessment criteria were largely met, however, few studies controlled for infant sex and age at assessment (Figure 3A). As 11 of the 15 cohort studies reported on cross-sectional neurodevelopmental measures, evaluating adequacy of follow up cohorts was often not applicable. The case series under review varied in quality, largely in terms of adequacy of participant characteristics, multiple-centre case collection, and clear and appropriate eligibility criteria (Figure 3B). High risk of performance and detection bias was detected in one RCT^52^, as it was not possible to blind participants or assessors to the study intervention, given the nature of the treatment (Figure 3C).

**Figure 3.**
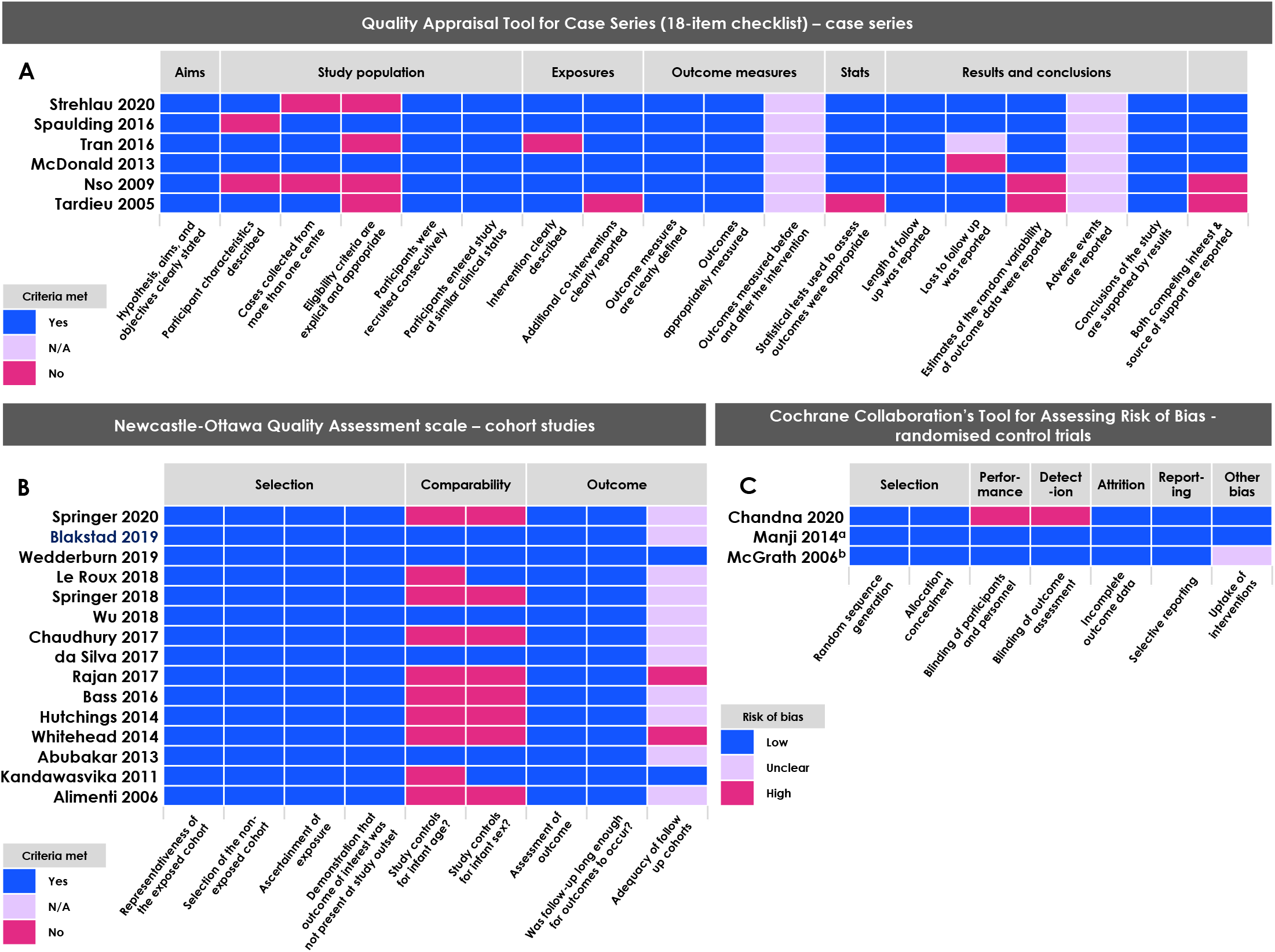
Quality assessment of articles according to study design using the (A) Newcastle-Ottawa Quality Assessment scale (cohort studies^23^), the Quality Appraisal Tool for Case Series (18-item checklist, case series^24^) and the Cochrane Collaboration’s Tool for Assessing Risk of Bias (randomized controlled trials^25^). ^a^This sub-study was a secondary endpoint of a larger randomised controlled trial. This sample includes children who attended only one of the three research clinics and 12% of the infants who attended follow up at 15 months overall (71.9% of the total number of infants who were randomized). Demographic characteristics across the placebo and multivitamin sub-study groups remained similar, so risk of selection bias (internal) was assessed as low. ^b^While authors report treatment compliance for the randomized arms, it is not clear that authors considered treatment compliance for the population of mothers whose infants had at least one neurodevelopmental assessment and were included in this analysis (327 of 1078 assigned to treatment arms).

### Infant HIV exposure status associates with neurodevelopmental outcomes in the first 36 months of life

A summary of study characteristics and key findings on neurodevelopmental outcomes from birth to 36 months of age in infants bom to mothers living with HIV is presented in Table 1.

### Cognitive outcomes

Infants who were HEI frequently scored lower than their HEU counterparts in measures of cognitive development before three years of age^36,40,45^ and were twice as likely to exhibit high risk for neurodevelopmental impairment when compared to HEU and HUU infant groups^38^. Reports on cognitive development for infants who were HEU (in comparison to HUU) were often inconsistent, with studies reporting associations between HEU and lower scores on measures of cognitive development^41,42^, as well as no difference in scores on measures of cognitive outcomes^49,50,53^. One study reported lower cognitive developmental scores for infants who were HEU compared to HUU, however, these differences did not persist after controlling for maternal substance use^47^. Cognitive developmental delay was often more prevalent among infants who were HEU at 12-13 months^49,50^ compared to HUU, but may not persist to three years of age^53^. One study reported that in comparison to reference norms, a higher proportion of infants who were HEU had average, higher average and superior performance on measures of cognitive outcomes^54^. Overall, HEU had a medium, negative effect on BSID-III cognitive subscale scores from four studies (7 cohorts; birth to 36 months) in comparison to HUU, however, the effect was not significant (−0.47 [-1.10, 0.15]; Figure 4). A funnel plot illustrating the scatter of effect estimates and standard error for the impact of HEU on infant cognitive outcomes is presented in Supplementary Figure 4. While the symmetry of the funnel suggests a possible negative skew, which could be a result of reporting bias, no test was performed to assess funnel plot asymmetry, as this is not recommended for meta-analyses with less than 10 studies^62^.

**Figure 4.**
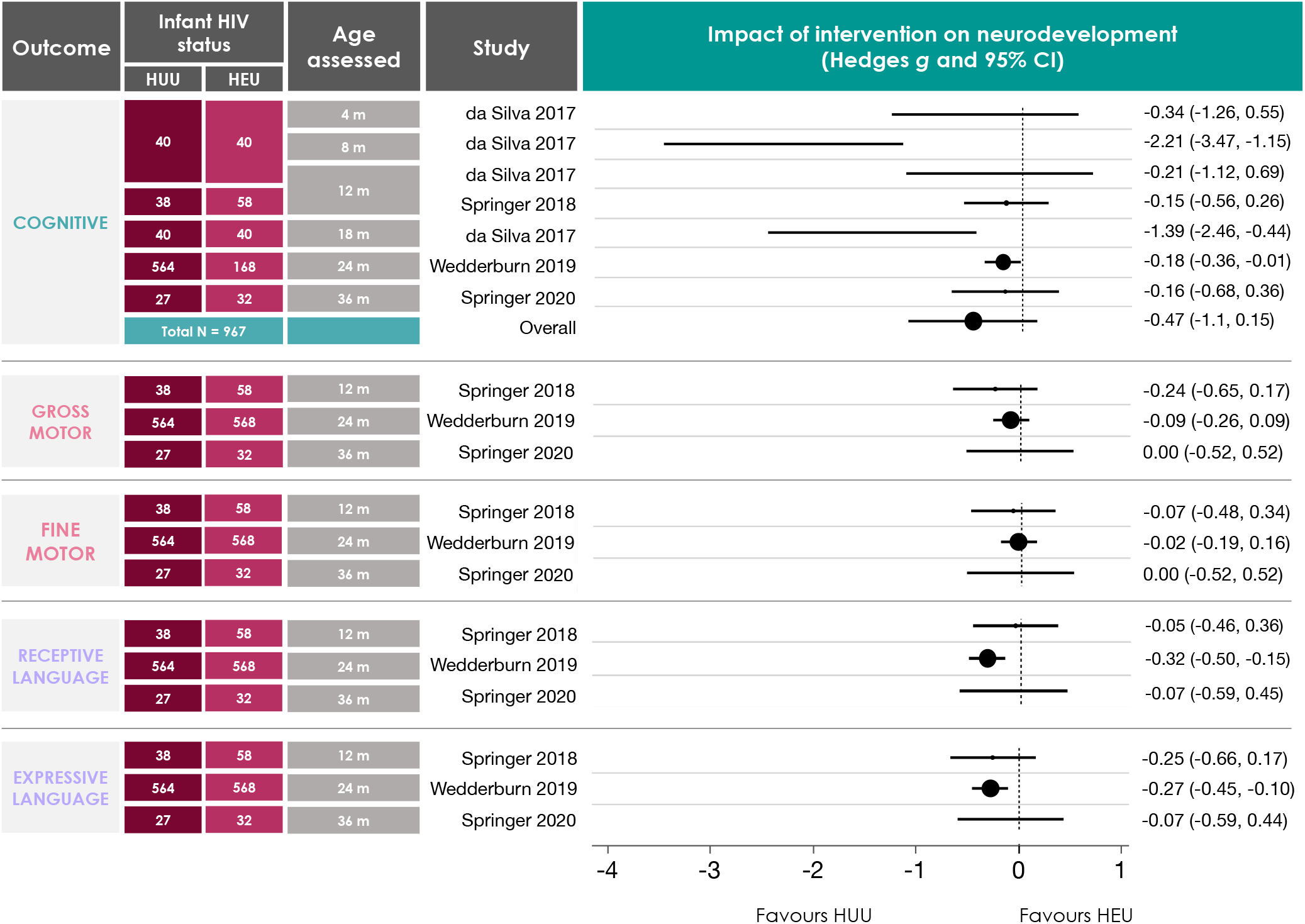
Random-effects meta-analysis for studies reporting on BSID-III sub-scales for infants who are HEU compared to HUU. Studies that reported means and standard deviations for scaled BSID-III scores were included. Where longitudinal data were available, analyses used the oldest data reported to capture any persistent impacts of HEU on neurodevelopment^50^. Data from da Silva et al. (2017) were cross-section for four separate groups of infants (aged 4, 8, 12 and 18 m). For one study, cognitive data analysed were composite scores, as scaled scores for this domain were not available^49^. Methodological quality assessments for these studies are reported in Figure 3. A summary effect estimate is only included for the cognitive sub-scale, as this is the only assessment that included more than five studies^31^. Data are presented in forest plots as Hedge’s ***g*** (95% Cl) in ascending order according to infant age at assessment. HUU = HIV-unexposed, uninfected; HEU = HIV-exposed, uninfected; m = months; BSID-III = Bayley Scales of Infant Development-3^rd^ edition^29^; CI = confidence interval.

### Motor outcomes

HEI often associated with lower scores on measures of motor development in comparison to infants who were HEU (up to 35 months of age^36,40,45,46^) and increased likelihood of motor developmental impairment compared to HEU and HUU infant groups^38^. While some studies suggested that infants who are HEU may also experience poorer motor outcomes in comparison to HUU^41,42^, others reported no differences in average motor scores between the two groups^47,49,50,53,50^ or evidence of motor delay for infants who are HEU^54,50^. One study reported psychomotor developmental delay in 3.64% of their sample (206 HEU) between birth and 18 months in comparison to a cited population rate of 1.1-2.5%^39^. Overall, HEU did not have a significant effect on BSID-III tine (−0.02 [-0.08, 0.04]) or gross (−0.10 [-0.29, 0.09]) motor subscale scores in three studies (Figure 4) from birth to 36 months, in comparison to HUU.

### Language outcomes

Of the three studies that reported on language outcomes in HEI infant populations, two reported lower scores^36,45^ in comparison to HEU, and one reported no differences^43^. Similar scores between HEU and HUU infant groups were reported for language assessments^53,56^ with no evidence of language delay for infants who are HEU^54^. One study reported lower scores and higher odds of delayed development in expressive and receptive language domains at 24 months (but not 6 months) among infants who were HEU compared to HUU in unadjusted and adjusted analyses^50^. Chaudhury et. al. also reported lower expressive language scores in infants who were HEU compared to HUU at 24 months, however, the differences were attenuated in adjusted analyses^57^. Overall, HEU had a significant and negative effect on expressive (−0.25 [-43, -0.08]), but not receptive (−0.25 [-0.61, 0.10]), language outcomes in comparison to HUU infants, in the first 36 months of life (Figure 4). Importantly, this combined effect estimate largely favours the results of one study, which had a much larger sample size (HUU: n=564, HEU: n=568)^50^ than the other two studies (HUU: n=38, HEU: n=58^49^; HUU: n=27, HEU: n=32 ^53^) that were included in this assessment.

### Behaviour outcomes

None of the studies under review assessed behavioural outcomes for infants who were HEI. Overall behavioural scores at 12 months^49^ and measures of communication, daily living and socialization from 18-36 months^47^ did not differ between infants who were HEU compared to HUU, however, a higher proportion of HEU infants (12.1 vs. 2.6% HUU) were classified as socially withdrawn^49^. In comparison to HUU infants, one study reported that infants who were HEU had higher scores in measures of personal-social development at 24 months^57^, while another reported lower adaptive behaviour scores for infants who were HEU between birth and 35 months^42^.

### Neurostructural outcomes

Three studies reported on neurostructural development in infants who were HEU^34,44,48^, one of which had a HUU comparison group^44^. Microcephaly, defined as a head circumference less than two standard deviations below the average^61^, and a risk factor for poorer neurodevelopmental outcomes until at least five years of age^63^, was recorded in 7.5% of infants who were HEU (105 of 1400) in one study that reported on data from six countries in Latin America and the Caribbean^34^. At least one neurologic condition (unspecified), was recorded in 9.6% of infants from these cohorts^34^. Of the two studies that characterised neural white matter in infants who were HEU, one recorded high prevalence of diffuse hyperintensity in the tegmentum pons and the supratentorial white matter^48^, and one recorded higher fractional anisotropy in the middle cerebellar peduncle region in infants who were HEU compared to HUU^44^. Associations between white matter structural signatures and performance on neurological assessments were also reported^44^.

### Relationships behveen early life nutritional factors and neurodevelopmental outcomes in infants perinatally exposed to maternal HIV infection and ART

A summary of study characteristics and key findings on the influence of nutrition-related factors and interventions on neurodevelopmental outcomes in infants born to mothers living with HIV is presented in Table 2. Infants who were HEU were breastfed at lower rates^57^ and for shorter durations^50,56^. Some studies report no associations between breastfeeding practices and neurodevelopmental outcomes up to 36 months^50,53,54^ or risk of developmental delay at 13 months^50^ in infants who were HEU. One study reported that HEU infants who were ever breastfed had higher cognitive and expressive language scores at 24 months when compared to infants who were never breastfed^57^. Notably, this study also reported associations between household food insecurity and lower gross motor scores, irrespective of maternal HIV status, however, higher rates of food insecurity were reported among infants who were HEU.

Three of the studies included were RCTs that aimed to evaluate the effects of a nutrition-related intervention on neurodevelopmental outcomes in infants exposed to maternal HIV infection and ART^51,52,58^. The interventions are described in full in Supplementary Table 3. Daily maternal multivitamin supplementation (from enrollment at 12-27 weeks’ gestation to 18 months postpartum) associated with higher scores on measures of motor development for infants at 6 months of age, an average increase in motor score of 2.6 points over the 6-18 month period, and reduced risk of motor developmental delay^51^. Daily infant multivitamin supplementation (from 6 weeks to 24 months postpartum) did not associate with any changes in performance on neurodevelopmental assessments performed at 15 months of age^58^. Notably, the mothers of all infants in this cohort also received daily multivitamin supplementation from enrollment to follow up^58^. Infants who received the Infant and Young Child Feeding (IYCF) intervention, which included maternal education on the importance of nutrition for infant health and development and a daily nutrient supplement given to infants from 6 to 18 months postpartum, did not have significantly improved neurodevelopmental outcomes compared to infants who received Standard-of-Care (SOC; Supplementary Table 3, Figure 5)^52^. However, when the IYCF intervention was given in conjunction with a Water, Sanitation and Hygiene (WASH) intervention (Supplementary Table 3), infants had higher gross and fine motor, language and behavioural outcomes at 24 months (Figure 5).

**Figure 5.**
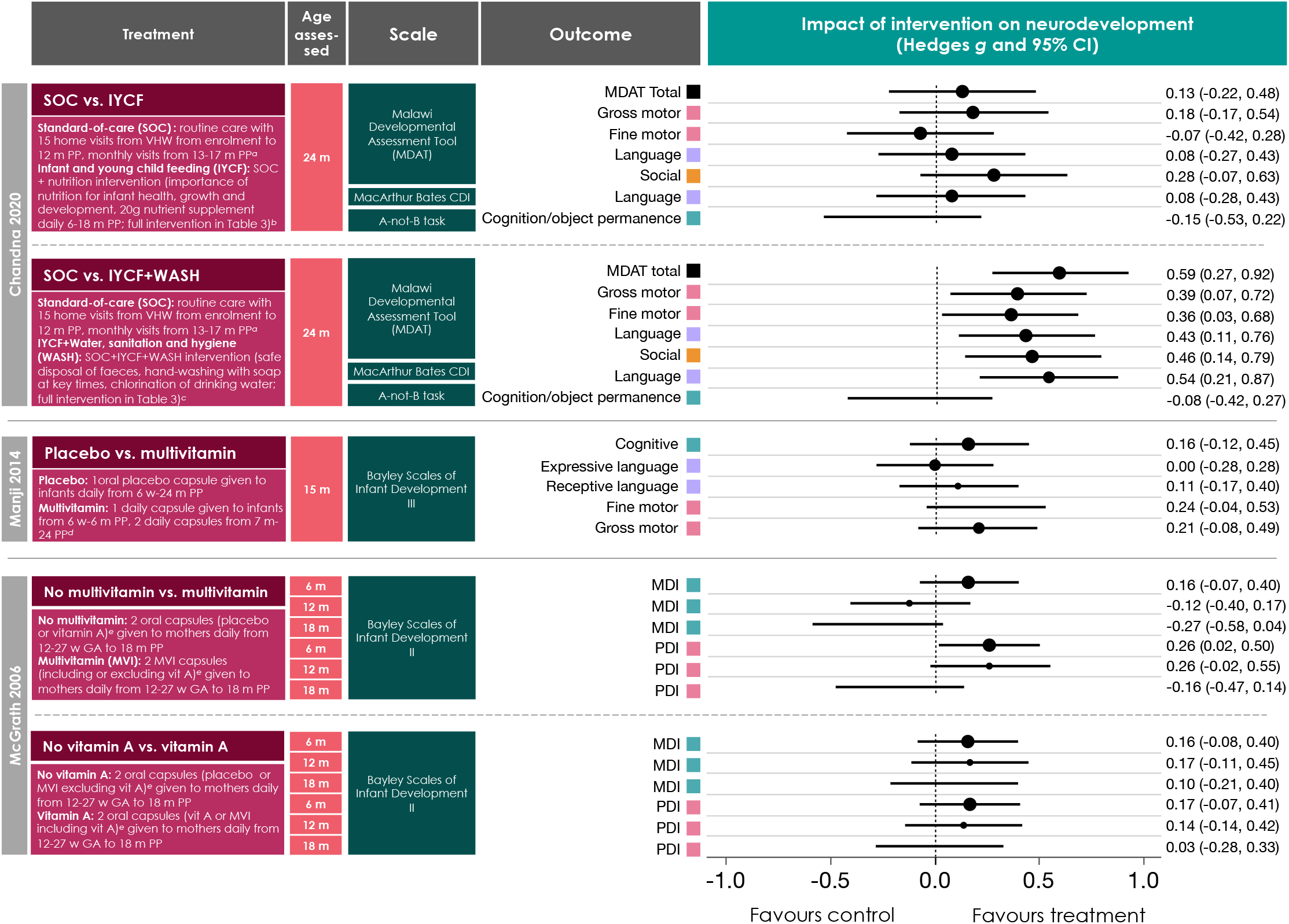
Summary of the results from randomised controls trials investigating the influence of early-life nutrition related interventions on neurodevelopmental outcomes in the first 24 m of life in infants exposed to HIV. Full descriptions of the study treatments or interventions are presented elsewhere (Supplementary table 3). Chanda *et al*. (2020) report positive effects of a combined IYCF+WASH intervention on motor, language and behavioural outcomes at 24 months in a group of 300 infants who are HEU. McGrath (2006) report positive effects of daily maternal multivitamin supplementation from 12-27 weeks’ gestation to 18 months postpartum on motor developmental outcomes in 327 infants exposed to maternal HIV infant (breakdown of infant HIV status not provided) at six months of age. Methodological quality assessments for these studies are reported in Figure 3. Data are presented in forest plots as Hedge’s *g* (95% CI) in ascending order according to infant age at assessment. SOC = Standard-of-care; ICYF = Infant and young child feeding; WASH = Water, sanitation and hygiene; VHW = Village health worker; MDAT = Malawi Developmental Assessment Tool; CDI = Communicative Development Inventories; d = days; w = weeks; m = months; PDI = Psychomotor development index; MDI = Mental development index; CI = confidence interval; HEU = HIV-exposed, uninfected.

### Sex differences and neurodevelopmental outcomes in infants perinatally exposed to maternal HIV infection and ART

Few studies ran analyses to examine differences in neurodevelopment for male and female infants. One study reported higher prevalence of ‘socially withdrawn’ classification among female HEU infants (87%) compared to male HEU infants^49^, however, the sample size was small (n=8). Female infants also had higher receptive communication scores than male infants in one population of infants who were HEU^54^. Male infants who were HEU were also more likely to have microcephaly or a diagnosed neurologic condition (not specified) between birth and six months^34^. For female infants who were HEU, the IYCF+WASH intervention associated with higher motor, language and social scores compared with standard-of-care, while male infants in the IYCF+WASH aim had higher language and social, but not motor scores, compared to SOC. There were no sex-differences for effect of infant multivitamin supplementation on neurodevelopmental outcomes at 15 months^58^.

## Discussion

In this formal evidence assessment and meta-analysis, we found that infants who were HEI had poorer neurodevelopmental outcomes in the first three years of life in multiple domains in comparison to infants who were HEU or HUU, which is in agreement with previous systematic reviews^12^’^64^. HEU appears to have persistent, negative effects on neurodevelopment in the first eight years of life^14^, albeit to a lesser extent than HEI, and our findings suggest that these negative effects may be detectable within the first three years of life. Further, maternal micronutrient supplementation from 12-27 weeks’ gestation to 18 months postpartum improved motor developmental outcomes at six months in infants exposed to maternal HIV infection, and a comprehensive nutrition intervention, encompassing educational measures and direct nutrient supplementation to infants who were HEU from 6 to 18 months postpartum, improved neurodevelopment across domains when given in conjunction with a WASH intervention^52^. Together, these findings suggest that the perinatal nutritional environment is a modifiable factor that can be optimised to improve child neurodevelopmental outcomes, which has important implications for the rising population of infants who are born HEU^65^.

Notably, reports of neurodevelopmental outcomes for infants who were HEU were inconsistent in their findings and are important to discuss within their respective contexts. Five studies that reported little or no differences between HEU and HUU infant groups across multiple domains were all South African cohorts^49,50,53,54,56^. South Africa faces the largest burden of HIV worldwide and has the largest treatment programme globally^66^. In 2018, it was estimated that 87% of women living with HIV who became pregnant had access to ART for PMTCT^67^. The extensive reach of PMTCT efforts in South Africa may contribute to the promising neurodevelopmental outcomes reported for infants who are HEU. How these findings may translate in different contexts, such as Brazil or Botswana, where two studies under review found poorer neurodevelopmental outcomes for infants who are HEU compared to HUU^41,57^, remains to be fully understood. Additionally, of the four studies that reported scaled or composite BSID-III scores and were summarised in Figure 4, all but one reporting on cognitive outcomes^41^, and all reporting on fine and gross motor, and receptive and expressive language outcomes were South African cohorts^49,50,53^. Notably, there may still be an overall negative effect of HEU on expressive language amongst South African cohorts^49,50,68^, suggesting that HEU may disproportionately affect language development even where PMTCT efforts are extensive, and additional effects to support language developmental trajectories may be necessary.

In studies reporting on sex differences in neurodevelopmental outcomes for infants who are HEU, male infants had poorer performance on measures of receptive communication^54^ and were more likely to have microcephaly or a diagnosed neurologic condition between birth a six months^34^. Further, the IYCF+WASH intervention improved motor scores for female, but not male, infants who were HEU^52^. Importantly, few studies overall reported sex-comparisons, and few cohort studies controlled for infant sex in analyses when comparing infant groups. The increased vulnerability of neurodevelopment in male infants when exposed to inflammatory or infectious factors is consistent with previous research^69,70^. As there are notable sex-differences in early brain development^71^, and sex-specific differences in breastmilk composition^72,73^ and breastfeeding practices^74,75^, it is critical that future studies examining relationships between HEU, early life nutrition and neurodevelopmental outcomes perform sex-based comparisons, present findings stratified by sex, and consider the presence of sex-dependent confounding. Doing so will fill a knowledge gap in whether, and to what extent, the effects of HEU may differentially affect male and female infants.

Notably, two of the studies investigating nutrition-related interventions reported promising effects for infants who are HIV-exposed^51,52^. While infant daily multivitamin supplementation from six weeks to 24 months showed no difference to the placebo group for neurodevelopmental outcomes in one cohort of infants who were HEU^58^, all the mothers in this cohort also received daily multivitamin supplementation from enrolment through follow up, which may have improved neurodevelopmental outcomes for this infant group overall. Unexpectedly, only one study reported beneficial associations between breastfeeding and neurodevelopment at 24 months^57^, irrespective of infant HIV exposure status, while others reported no associations between breastfeeding practices and neurodevelopment in HEU and HUU infant groups. However, detailed data on length of exclusive breastfeeding^53,56^ or mixed feeding^54^ was often not reported, limiting our ability to draw conclusions.

The lack of available data on maternal nutrient intakes or levels is a key limitation in the studies reviewed and an opportunity for future research. Maternal nutrient status during pregnancy and the postpartum period is critical to supporting rapid growth and development of the infant^76^, and maternal diet in part determines the nutritional composition of breastmilk^77^. Further, how maternal HIV infection and ART may alter breastmilk composition is not well understood. Investigation of breastmilk immune factors among women living with HIV remains limited, and among the few studies that have measured these factors, varied results have been reported. Higher levels of non-specific IgA^78^ and IgG^78,79^ have been measured in breastmilk from women living with HIV, and higher levels of fucosylated human milk oligosaccharides in breast milk are associated with lower infant mortality among infants who are HEU, but not HEI, during breastfeeding^80^. There is opportunity to better understand how the benefits of exclusive breastfeeding, including improved maternal health outcomes^81^ and constituents in breastmilk^82^, could be protective against any adverse effects HEI or HEU on infant neurodevelopment.

Importantly, there are key variables that interact to influence fetal and infant development in pregnancies complicated by maternal HIV infection that were beyond the scope of this review and are important to consider. We did not evaluate relationships between maternal or infant ART and infant neurodevelopment. Previous research has shown that the timing and type of ART initiation for infants who are HIV-exposed may affect neurodevelopmental outcomes^83,84^. Further, as vulnerabilities of the developing fetal brain to the effects of infectious exposures and inflammation temporally vary throughout pregnancy^85,86^ the timing of maternal HIV infection, viral suppression and related treatments is an important consideration and these data were not frequently available. In 2017, one third of newly transmitted HIV infections in the Middle East and North Africa, eastern Europe and central Asia occurred in people who injected drugs^65^, and neurodevelopmental risks for the fetus and infant related to these comorbid exposures *in utero* are important to understand. Of the studies assessed here, one reported that maternal substance use in pregnancy had a greater (negative) effect on neurodevelopmental trajectories than infant HIV and ART exposure status^47^. Low socioeconomic status is also a known risk factor for poorer cognitive development^87^, and is particularly important to consider in the context of HIV infection, given the overlaps between HIV/AIDS and economic and food insecurity^88^. Lastly, poor maternal mental health during the pre- and postnatal periods may adversely influence infant neurodevelopmental outcomes^89^, and among women living with HIV specifically, depression and mental health vulnerabilities are especially prevalent^90^ and have shown to associate with infant health outcomes^91^.

Some maternal ART are also associated with increased risk of preterm birth (<37 weeks’ gestation)^92^, and preterm birth has independent consequences for infant neurodevelopment^93^. Here, preterm birth was a risk factor for lower cognitive and gross motor scores among infants who were HIV-exposed in one study^55^, and among 1400 infants who were HEU in another study^34^, those who had diagnosed microcephaly or a neurological condition in the first 6 months of life were more likely to have a lower gestational age at birth. Thus, it is critical to consider how premature birth and *in utero* exposure to maternal HIV/ART may have additive, negative effects for offspring neurodevelopment. Notably, risk of premature delivery among women living with HIV on ART is exacerbated by poor maternal nutritional status^92^. This suggests that interventions to target prenatal maternal nutritional status could improve neurodevelopment in infants who are HIV-exposed through both a reduction in premature birth prevalence, and by the direct effects of improved nutrient resources to support neurodevelopmental processes.

While our findings suggest that the early nutritional environment may be leveraged to improve early development and health outcomes before three years of age for the rising number of infants who are born HEU, our ability to draw conclusive statements from this review is limited by the small quantity of published research on this topic. Limiting our inclusion criteria to articles published following the launch of international PMTCT efforts likely contributed to the low number of included articles, however, this was necessary for ensuring comparability and translatability to current day contexts. Further research on how early life nutritional exposures can be enhanced to improve health outcomes for infants who have been exposed to HIV is needed and would allow us to tailor nutrition-related interventions during critical periods of development to match their specific needs. Additionally, our analysis of reported associations between white matter structural abnormalities, in brain regions involved in memory, learning, emotional and cognitive processing^94,95^, and performance on neurodevelopmental assessments as early as two to four weeks postpartum, suggests an opportunity for early neurodevelopmental screening to identify infants who may be susceptible to suboptimal developmental outcomes^44^. An ability to predict, at an early stage, which infants who have been exposed to HIV are at risk of poorer neurodevelopmental trajectories, together with a clear understanding of how the early nutritional environment may be optimised for mothers living with HIV and their infants, are key for determining which infants will benefit from additional support during critical developmental periods in order to mitigate adverse outcome later in life.

## Data Availability

Not Applicable

## Supplementary figures

**Supplementary Figure 1.**
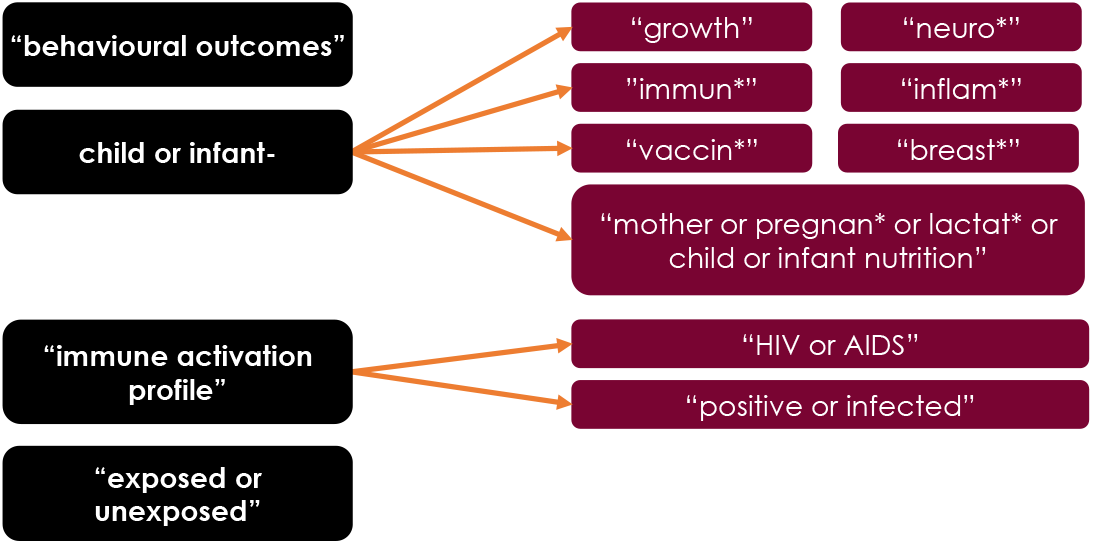
Evidence-based review keyword search terms.

**Supplementary Figure 2.**
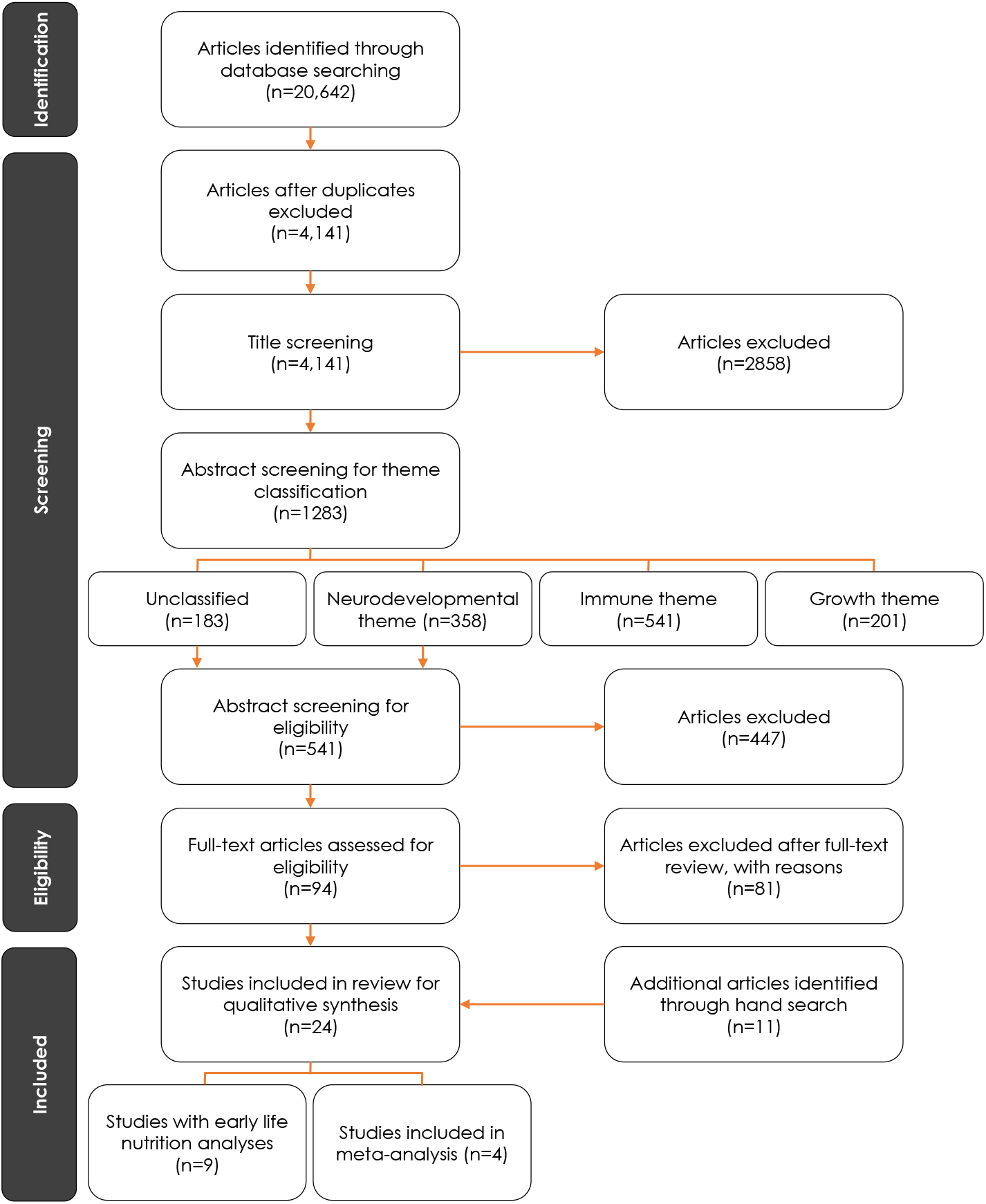
PRISMA flow diagram for article selection. Additional search by hand was performed on March 25, 2020 using the same set of pre-determined key words in PubMed, CINAHL, ProQuest, and Web of Science.

**Supplementary Figure 3.**
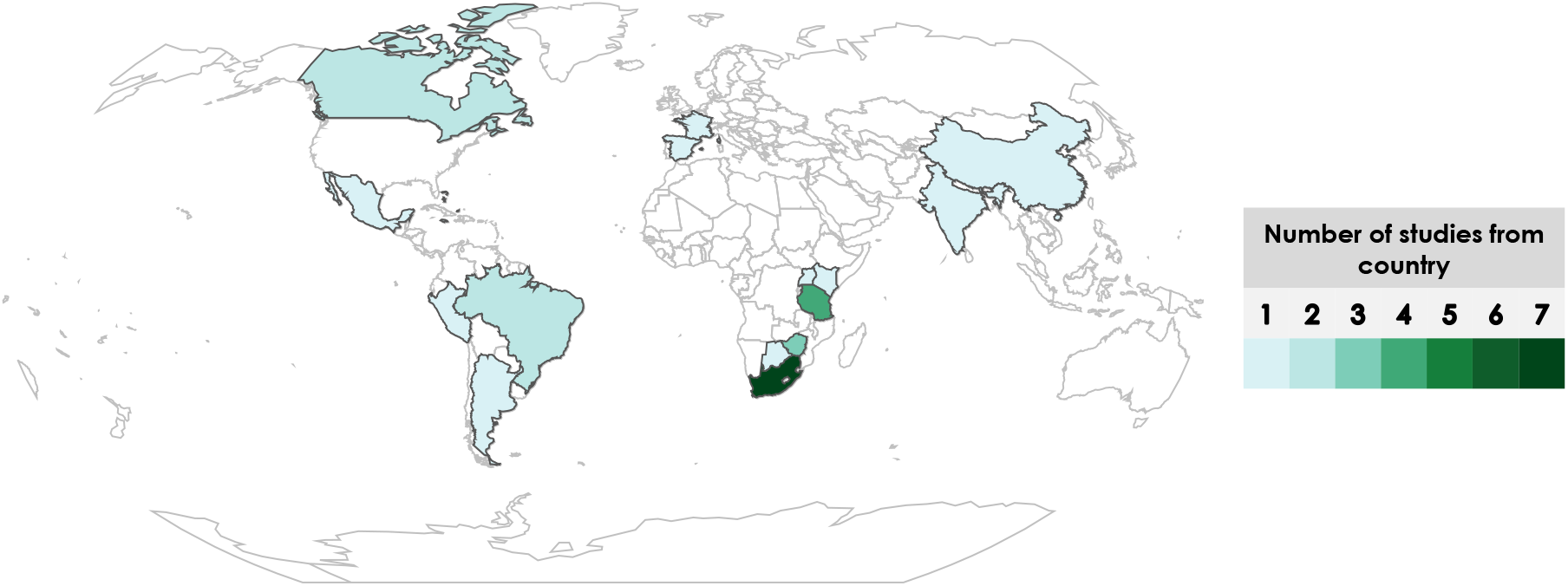
Locations of studies considered in review. South Africa (n=7) had the highest representation in studies under review. 57% (n=17) of the cohorts included in studies under review were from Africa, followed by 17% (n=5) from North America, 13% (n=4) from South America, and 7% (n=2) from both Asia and Europe. One study report on data from cohorts in Brazil, Argentina, Peru, Mexico, Bahamas, and Jamaica (Spaulding et al., 2016). For the purpose of this figure, each of these cohorts was counted once.

**Supplementary Figure 4.**
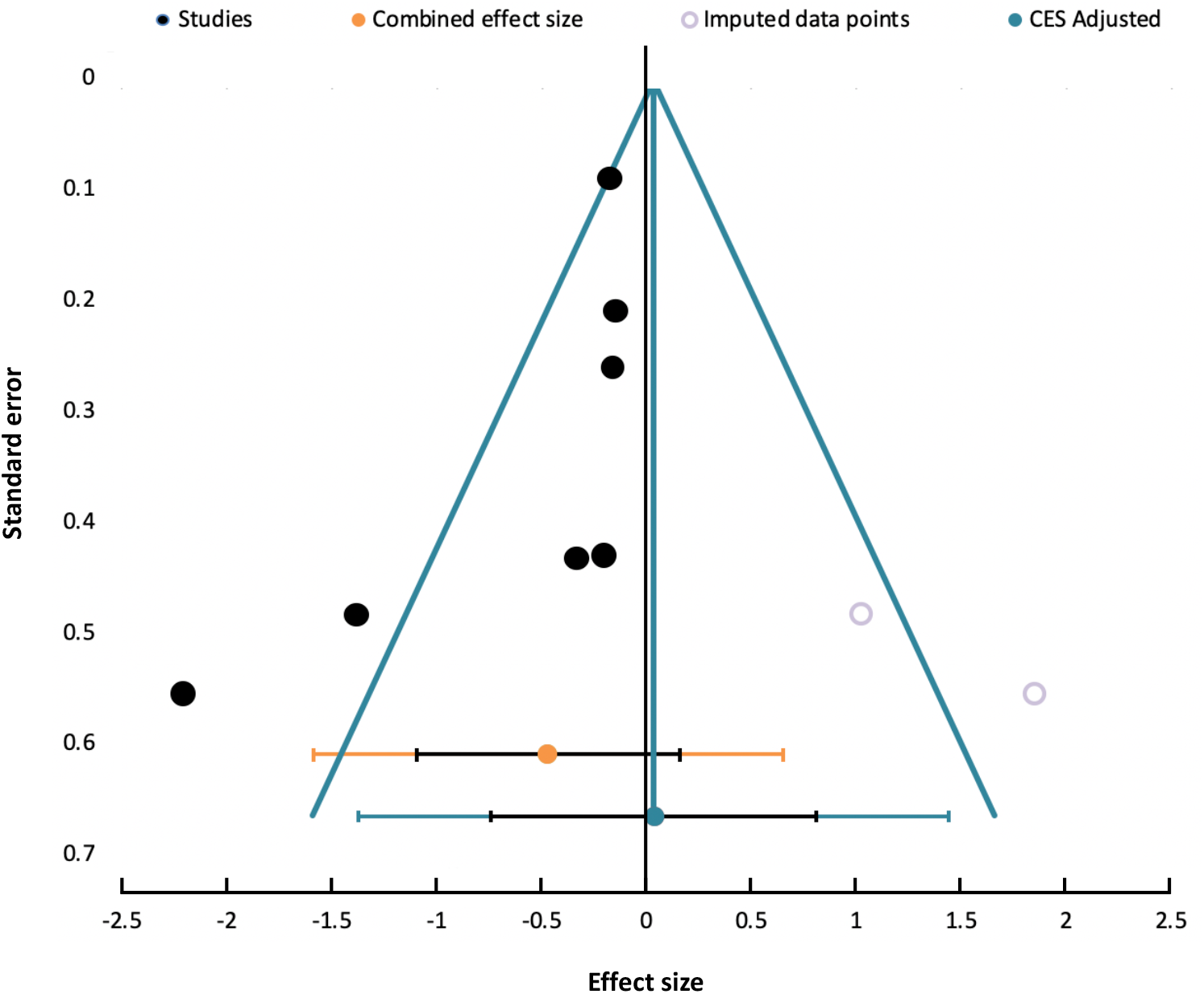
Funnel plot with effect estimates and standard error for meta-analysis of cognitive sub-scale composite and scaled scores (Bayley Scales of Infant Development, 3^rd^ ed.) from four studies (7 cohorts).

**Supplementary Table 1.**
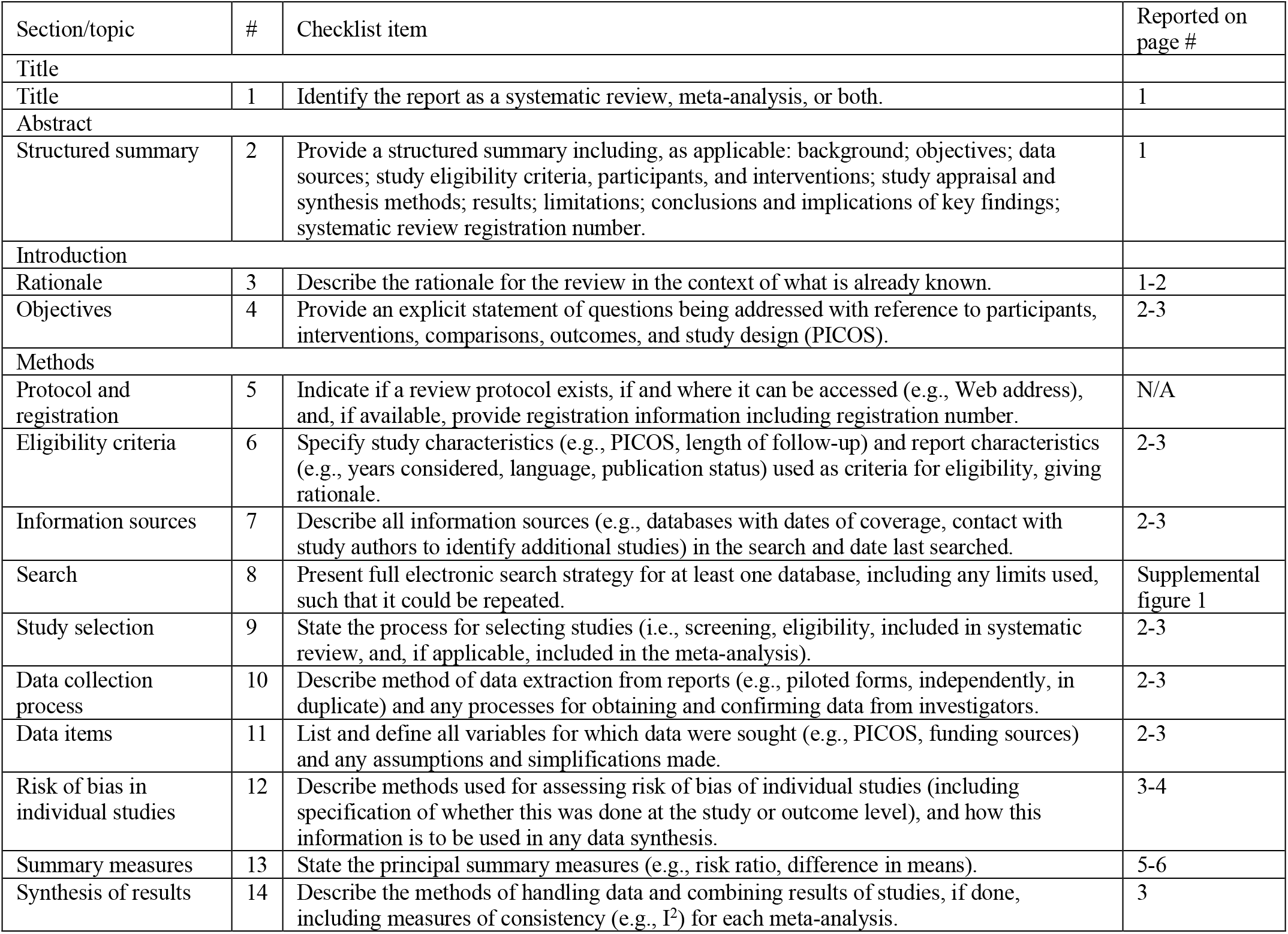
PRISMA Checklist^19^.

| Section/topic | # | Checklist item | Reported on page # |
| --- | --- | --- | --- |
| Title |  |  |  |
| Title | 1 | Identify the report as a systematic review, meta-analysis, or both. | 1 |
| Abstract |  |  |  |
| Structured summary | 2 | Provide a structured summary including, as applicable: background; objectives; data sources; study eligibility criteria, participants, and interventions; study appraisal and synthesis methods; results; limitations; conclusions and implications of key findings; systematic review registration number. | 1 |
| Introduction |  |  |  |
| Rationale | 3 | Describe the rationale for the review in the context of what is already known. | 1-2 |
| Objectives | 4 | Provide an explicit statement of questions being addressed with reference to participants, interventions, comparisons, outcomes, and study design (PICOS). | 2-3 |
| Methods |  |  |  |
| Protocol and registration | 5 | Indicate if a review protocol exists, if and where it can be accessed (e.g., Web address), and, if available, provide registration information including registration number. | N/A |
| Eligibility criteria | 6 | Specify study characteristics (e.g., PICOS, length of follow-up) and report characteristics (e.g., years considered, language, publication status) used as criteria for eligibility, giving rationale. | 2-3 |
| Information sources | 7 | Describe all information sources (e.g., databases with dates of coverage, contact with study authors to identify additional studies) in the search and date last searched. | 2-3 |
| Search | 8 | Present full electronic search strategy for at least one database, including any limits used, such that it could be repeated. | Supplemental figure 1 |
| Study selection | 9 | State the process for selecting studies (i.e., screening, eligibility, included in systematic review, and, if applicable, included in the meta-analysis). | 2-3 |
| Data collection process | 10 | Describe method of data extraction from reports (e.g., piloted forms, independently, in duplicate) and any processes for obtaining and confirming data from investigators. | 2-3 |
| Data items | 11 | List and define all variables for which data were sought (e.g., PICOS, funding sources) and any assumptions and simplifications made. | 2-3 |
| Risk of bias in individual studies | 12 | Describe methods used for assessing risk of bias of individual studies (including specification of whether this was done at the study or outcome level), and how this information is to be used in any data synthesis. | 3-4 |
| Summary measures | 13 | State the principal summary measures (e.g., risk ratio, difference in means). | 5-6 |
| Synthesis of results | 14 | Describe the methods of handling data and combining results of studies, if done, including measures of consistency (e.g., $I^2$) for each meta-analysis. | 3 |

**Supplementary Table 2.** Methodological quality assessment criteria set a priori.

| Checklist item requiring specification | Criterion |
| --- | --- |
| **Newcastle-Ottawa Quality Assessment scale (cohort studies)<sup>24</sup>** |  |
| Comparability | 1. Determined based on whether the authors controlled for infant sex and age at assessment in analyses |
| Was follow-up long enough for outcome to occur? | 1. As neurodevelopment within the first three years of life was the outcome of interest, demonstration that outcome of interest was not present at the start of the study was not required and all articles received a ‘yes’ assessment for this criterion. |
| Adequacy of follow up cohorts | 1. Where cohorts had cross-sectional neurodevelopmental data, adequacy of follow up cohorts was not assessed;<br>2. Where neurodevelopmental data were longitudinal, adequacy of follow up cohorts was considered where subjects lost to follow up were minimal (<20%) or analyses were run to establish similarity between infants retained at follow up vs. not. |
| **Quality Appraisal Tool for Case Series (18-item checklist)<sup>25</sup>** |  |
| Are characteristics of the participants included in the study described? | 1. Characteristics of the cohort that were important to include were pre-defined as: number of participants (infants), age range of infants with neurodevelopmental assessment and infant sex. |
| Did participants enter the study at a similar point in the disease? | 1. Based on our outcomes of interest, this was redefined as “Did participants enter the study at a similar point in development (i.e., age)”. |
| Was the intervention clearly described in the study? | 1. “Intervention” was modified to be “exposure” of interest, and defined as: Maternal HIV infection and information on ART;<br>2. A point was given if authors reported details on whether or not the mothers were on ART and what ART treatments mothers were on. |
| Were additional (co-interventions) clearly described in the study? | 1. “Co-intervention” was modified to be “co-exposure” of interest, and defined as: infant HIV exposure status and information on ART;<br>2. A point was given if authors reported details on whether or not an infant contracted HIV infection prior to or during the study, or received ART intervention |
| Are adverse events reported? | 1. Adverse events were not considered as no intervention was being employed. |
| **Cochrane Collaboration’s Tool for Assessing Risk of Bias<sup>26</sup>** |  |
| Other bias | 1. Defined as assessment of compliance to intervention |

**Supplementary Table 3.**
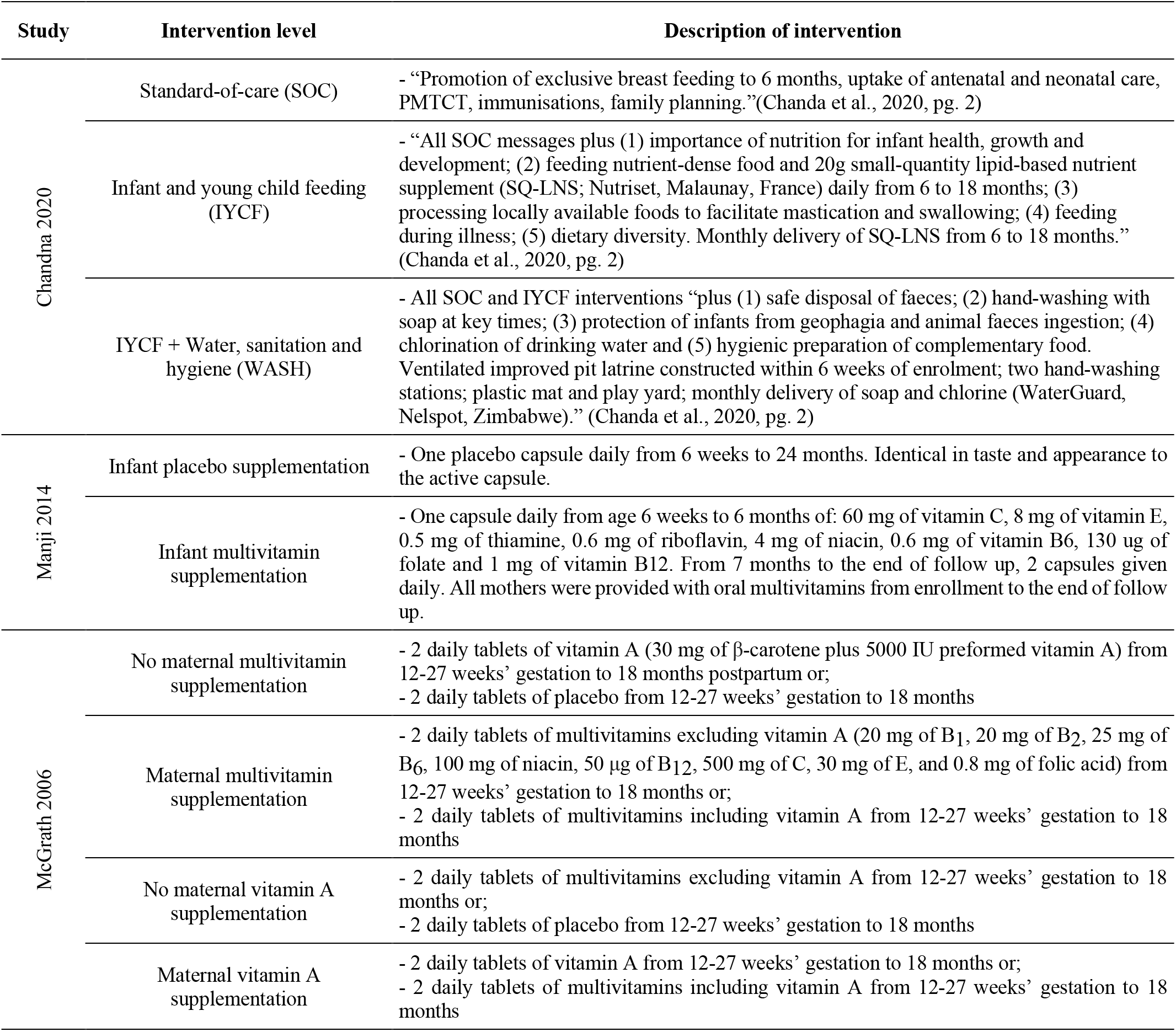
Descriptions of trial interventions for randomized control trials on early life nutrition-related factors and neurodevelopment in infants exposed to HIV.

